# Gene copy number variation in pediatric mental illness in a general population

**DOI:** 10.1101/2022.09.12.22279764

**Authors:** Mehdi Zarrei, Christie L. Burton, Worrawat Engchuan, Edward J Higginbotham, John Wei, Sabah Shaikh, Nicole M. Roslin, Jeffrey R. MacDonald, Giovanna Pellecchia, Thomas Nalpathamkalam, Sylvia Lamoureux, Roozbeh Manshaei, Jennifer Howe, Brett Trost, Bhooma Thiruvahindrapuram, Christian R. Marshall, Ryan K.C. Yuen, Richard F. Wintle, Lisa J Strug, Dimitri J. Stavropoulos, Jacob A.S. Vorstman, Paul Arnold, Daniele Merico, Marc Woodbury-Smith, Jennifer Crosbie, Russell Schachar, Stephen W. Scherer

**Author notes:** Correspondence: Stephen W. Scherer. These authors contributed equally (Mehdi Zarrei, Christie L. Burton, Worrawat Engchuan). These authors share senior authorship (Jennifer Crosbie, Russell Schachar, Stephen W. Scherer).

## Abstract

We assessed the relationship of gene copy number variation (CNV) in mental health/neurodevelopmental traits and diagnoses, physical health, and cognitive biomarkers in a community sample of 7,100 unrelated European, and East Asian children and youth (Spit for Science). Diagnoses of mental health disorders were found in 17.5% of participants and 27.6% scored in the highest 10% on either or both ADHD and OCD trait measures. Clinically relevant CNVs were present in 3.9% of participants and were associated with elevated scores on a continuous measure of ADHD (*p*=5.0×10^−3^), on a cognitive biomarker of mental health (response inhibition (*p*=1.0×10^−2^)), and on prevalence of mental disorders (*p*=1.9×10^−6^, odds ratio: 3.09). With a rise of mental illness, our data establishes a baseline for delineating genetic contributors in paediatric-onset conditions.

**One Sentence Summary:** Copy number variation predicts neurodevelopmental and mental health phenotypes in the general population.

---

Many children and youth suffer from impairing and persistent mental illnesses such as attention deficit/hyperactivity disorder (ADHD), obsessive-compulsive disorder (OCD) and autism spectrum disorder (ASD). Twin, genetic and molecular studies indicate that genetic risk factors along with environmental influences create variation in neurodevelopment with disorders representing the extremes of typically widely distributed cognitive and behavioral traits that underly mental health state.

Copy number variation (CNV) refers to deletions or duplications of segments of DNA that alter the typical diploid state of genes and/or their regulatory elements along chromosomes. CNV is ubiquitous in all genomes and when such genetic events (new large CNV arises ∼1/100 meioses) impact important brain developmental genes it can cause or increase risk to mental illness. Autism spectrum disorder (ASD) (*1*) and schizophrenia are the prototypical examples, but CNVs are also found involved in most every other brain condition analyzed (Supplemental Table S1). Some rare (<1% population frequency) CNVs are so strongly related to mental illness that they are recognized by the American College of Medical Genetics guidelines (*2*) as pathogenic or likely pathogenic CNVs (significant).

Most studies of the relationship between rare CNVs and mental health disorders are currently based on examining clinical databases of individuals with ASD, ADHD or intellectual deficiency. There have been few general population studies of the prevalence and mental health significance of CNVs in children. However, these general population studies have tended to focus on only a narrow range of mental health conditions, types of CNVs (typically large deletions), and have not distinguished among CNVs according to established risk (significant versus susceptibility). With increasing use of genome-wide analysis in clinical care (*3, 4*), we developed a study with the aim of advancing our understanding of CNV and the genes impacted by it on behavioral and cognitive traits.

We examined the prevalence of rare CNVs and their associated phenotypes in a large community-based sample of European and East Asian children and youth (N=7,100; Spit for Science study) (4-18 years of age; mean=10.7±2.9). Participants and their parents were visitors to a Science Centre. The sample had a male to female sex ratio of 1.01 (male: 3,559; female: 3,541; Table 1 and Supplementary Table S2). We focused on the two largest ancestry populations: European (n=5,686; M:F=1.04) and East Asian (n=1,414; M:F=0.88) for a total of 7,100 participants (7,050 with complete phenotype information). We obtained parent or self-reported information about the participants diagnoses of mental, neurodevelopmental and physical health and used reliable and valid rating scales to measure traits associated with ADHD (inattention and hyperactivity-impulsivity), OCD, anxiety, and ASD (see Supplementary Information). Participants performed cognitive biomarker task, the stop-signal task (SST), to measure the response inhibition (stop-signal reaction time - SSRT) and reaction time variability (RTV). Longer SSRT indicates worse inhibitory control and greater RTV indicates variability in performance. SSRT and RTV are putative cognitive biomarkers for mental health / neurodevelopmental (ND) disorders in childhood (*5, 6*). We categorized each participant as having a “high trait score” if their scores within the most extreme 10% of the sample as well as examining continuous trait measures.

**Table 1:** Rate of Mental Health/Neurodevelopmental Disorders. Distribution of inferred or self-reported diagnosis of neurodevelopmental or neuropsychiatric disorders or traits.

| Category | Phenotype | N | (%)<sup>§</sup> |
| --- | --- | --- | --- |
| Diagnosis | ADHD | 474 | (6.72%) |
|  | OCD | 80 | (1.13%) |
|  | ASD | 145 | (2.07%) |
|  | Anxiety | 320 | (4.54%) |
|  | Tics/Tourette's | 84 | (1.19%) |
|  | Learning Disorder/Learning Problem | 644 | (9.13%) |
|  | Intellectual Disability/Developmental Delay | 13 | (0.18%) |
|  | Mood Disorder | 96 | (1.36%) |
|  | Schizophrenia/Bipolar/Manic Depressive | 6 | (0.09%) |
|  | Language Disorder | 5 | (0.07%) |
|  | Motor Delay/Motor Disability | 4 | (0.06%) |
|  | Eating Disorder | 2 | (0.03%) |
| Any Diagnosis |  | 1,232 | (17.48%) |
| Any High Trait |  | 1,948 | (27.63%) |
| Any Diagnosis or High Trait |  | 2,412 | (34.21%) |

§Overall *n* used is restricted to those that have diagnosis and trait data (Total *n*=7,050, 5,660 of European Ancestry, and 1,390 of East Asian Ancestry).
Abbreviations: N: number of samples in each designated category; ADHD: attention deficit/hyperactivity disorder; SWAN: The Strengths and Weaknesses of Attention-Deficit/Hyperactivity Disorder Symptoms and Normal Behavior Scale; OCD: obsessive-compulsive disorder; TOCS: Toronto Obsessive-Compulsive Scale; ASD: autism spectrum disorder. (see Supplementary Text for details).
\*High Traits: Top 10% of SWAN (Either or total score, Inattentive and/or Hyperactive/Impulsive), TOCS, CBCL (Spit for Science 1) RCADS (Spit for Science 2), AQ (Spit for Science 2) t-scores. Total cases with each trait are as follows: ADHD (7,023), OCD (7,010), Anxiety (5,607), ASD (1,787).
†Only measured in Spit for Science 2

Diagnoses of mental health /ND disorders were reported in 17.5% (1,232/7,050) of participants and 27.6% (1,948/5,050) scored in the highest 10% on either or both ADHD and OCD trait measures (high trait) (Fig. 1A; Table 1 and Supplementary Table S2). Cumulatively, 34.2% (2,412/5,050) of participants reported either a mental health disorder or scored in the high trait group (Table1 and Supplementary Table S2) and 10.9% (768/7,050) had both. Learning disabilities/learning problems were the most frequently reported disorder (9.1%) followed by ADHD (6.7%); 15.1% of participants were classified as having high ADHD traits, 10.6% as high anxiety traits, 10% as high OCD and 7% as high ASD traits (Table 1).

**Fig. 1:**
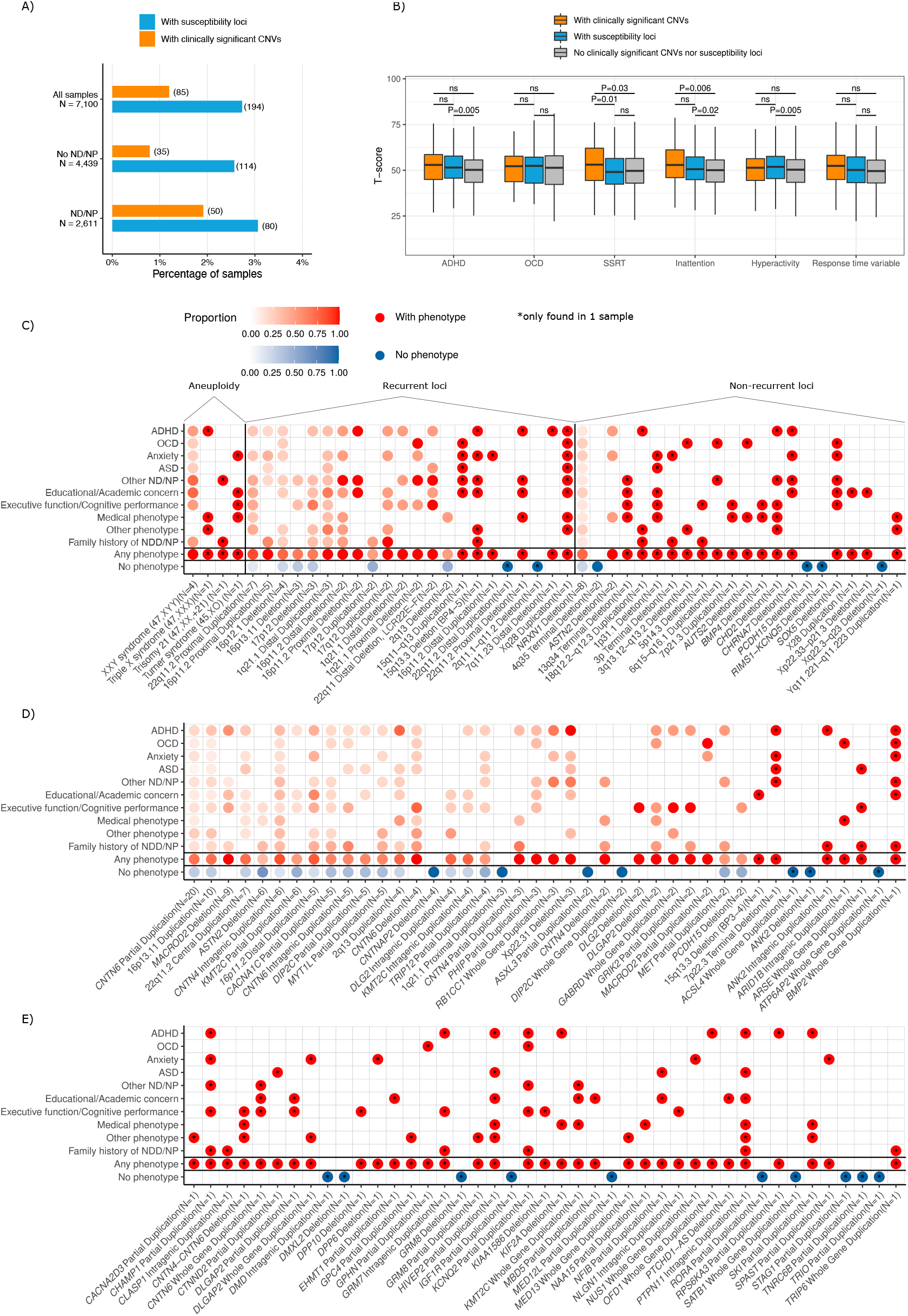
A) proportion of samples with a diagnosis of NDD/NP or traits, and the proportion of each category carrying clinically relevant CNVs, i.e. clinically significant CNVs or susceptibility CNVs, B) comparison of T-scores for OCD, ADHD, or stop-signal reaction-time (SSRT) and reaction time variability (RTV) traits between subjects carrying clinically significant CNVs, susceptibility CNVs, and those without clinically relevant CNVs, and C) distribution of self-reported or inferred diagnosis or traits of mental illness in participants carrying clinically significant CNVs, and D and E) distribution of self-reported or inferred diagnosis or traits of neurodevelopmental or neuropsychiatric disorders in participants carrying susceptibility CNVs. We confirmed the validity of these variants using qPCR or by visualizing the B-allele frequency. Diagnosis of mental health /ND disorders are based on self-reported information or inferred from high traits (top 10%). The number on the bars (panel A) indicates the number of participants carrying CNVs in each category. Abbreviations: ADHD: attention deficit-hyperactivity disorder; OCD: obsessive-compulsive disorder, ASD: autism spectrum disorder, ND: neurodevelopmental disorders, NP: neuropsychiatric disorders; SSRT: response inhibition using the Stop-Signal task. Other ND/NP: a reported diagnosis of Tics/Tourette’s, learning problem/disability, eating disorder, mood disorder, schizophrenia/bipolar/manic depressive, motor disability, intellectual disability/developmental delay, and/or language disorder (s). Medical Phenotype: a reported diagnosis or treatment of medical disorders and genetic conditions (e.g., epilepsy, Scoliosis, Diabetes, Ulcerative Colitis, 1q21.1 duplication). Other Phenotype: an additional phenotype not covered by the other categories that may be relevant to identified CNVs (e.g., premature birth, assisted reproductive technology for pregnancy, maternal infection during pregnancy). For details refer to Supplementary Table S3.

DNA was extracted from saliva samples and genotyping was performed on Illumina Infinium HumanCoreExome beadchips (5,220/7,100=73.5%) or Illumina Infinium Global Screening Array (1,888/7,100=26.5%; Supplementary Table S2). A high-quality set of CNVs was identified as those called by at least two of three algorithms (*7*). Variant size ranged from 10 kb to sex chromosome aneuploidies (n=7) and a trisomy of chromosome 21 (Table 1). The relevant genotype data and CNVs are being prepared for submission to the European Genome-Phenome Archive and the dbVar Database, respectively. Experimental details are available as Supplementary Materials.

Using criteria specified in Supplementary Materials, we identified 287 (3.9%) clinically relevant CNVs in 279 of 7,100 individuals across both European and East Asian cohorts (Tables 2; Supplementary Table S3; Fig. 1A). Of these, 1.2% (85/7,100) of participants harbored clinically significant CNVs and 2.7% (194/7,100) carried susceptibility CNVs. Participants of European (223/5,686=3.9%) and East Asian (56/1,414=3.6%) ancestries had similar rates of clinically relevant CNVs (*p*> 0.05), clinically significant and susceptibility CNVs (*p*> 0.05).

**Table 2:**
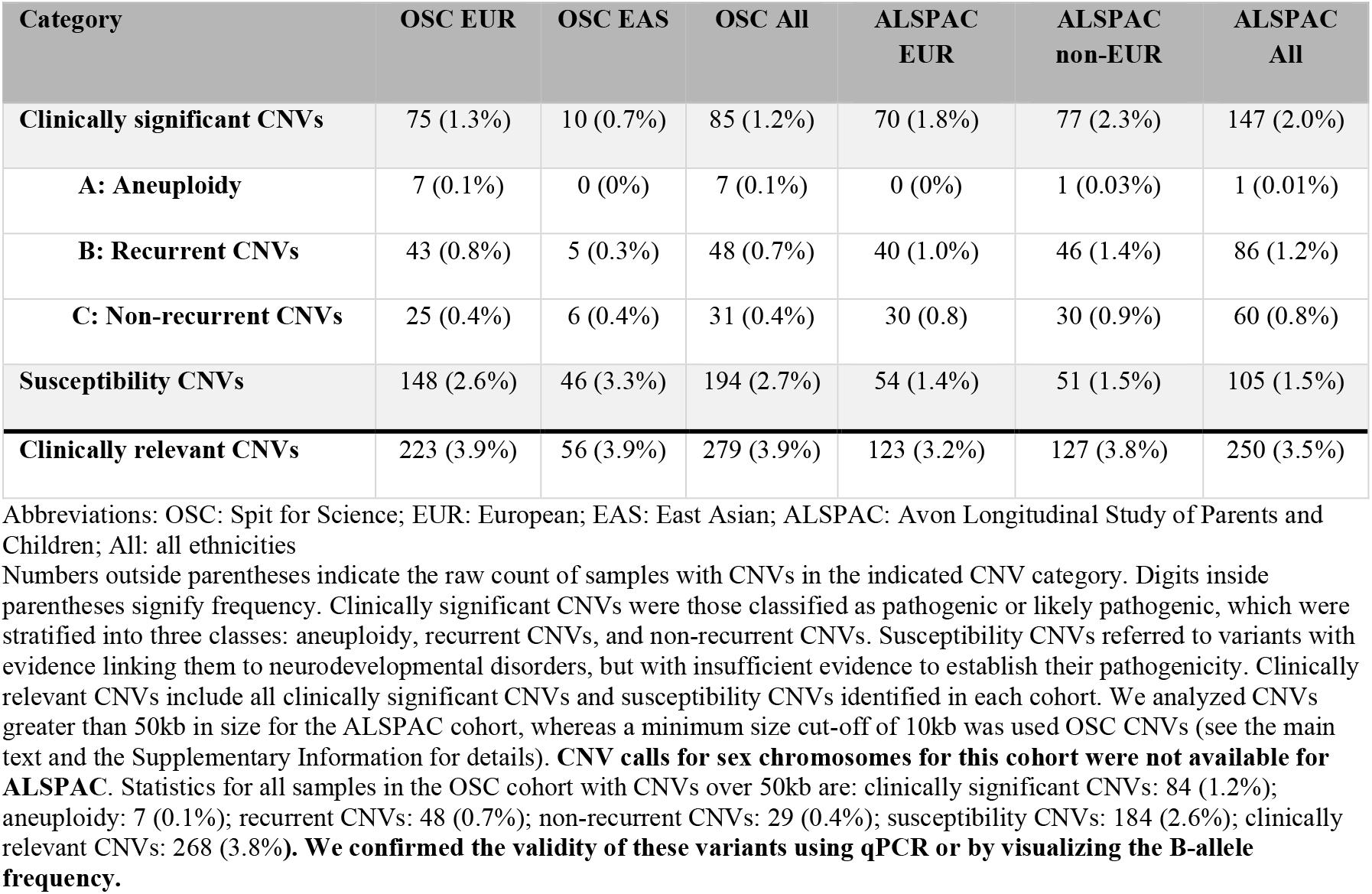
Distribution of different clinically relevant CNVs identified in 7,100 samples in Spit for Science (n=5,686 for European and 1,414 for East Asian) and 7,219 samples in Avon Longitudinal Study of Parents and Children (n=3,890 for European and 3,329 in non-European).

## Clinically relevant CNVs increase likelihood of mental health disorders or high traits

Of 279 participants with clinically relevant CNVs in Spit for Science cohort, 28% (78/279) reported a mental health/neurodevelopmental/neuropsychiatric disorder (NDD/NP), 38.7 % (108/279) met criteria for membership in one of the high trait groups, and 46.6 % (130/279) had either a diagnosis or high traits (Table 3; Supplementary Table S2). We found that 59% and 41% of participants with significant and susceptibility CNVs respectively had reported diagnosis or high traits (Table 3) compared to 34% of participants without either type of CNV.

**Table 3:** Enrichment of clinically significant, susceptibility, and either type of CNVs among participants with self-reported diagnoses of neurodevelopmental/neuropsychiatric (ND/NP) disorders or with ND/NP traits (n=7050).

| Category | Count | Reported Dx (*All ND/NP*)<sup>a</sup><br>(n=1232, 17%) |  |  | High Traits<sup>b</sup><br>(n=1948, 28%) |  |  | Reported Dx *and/or* High Traits<br>(n=2412, 34%) |  |  |
| --- | --- | --- | --- | --- | --- | --- | --- | --- | --- | --- |
|  |  | YES (%) | NO (%) | OR | YES (%) | NO (%) | OR | YES (%) | NO (%) | OR |
| Significant CNVs | 85 | 33 (39%) | 52 (61%) | 3.09**** | 39 (46%) | 46 (54%) | 2.27*** | 50 (59%) | 35 (41%) | 2.81**** |
| No significant or susceptibility CNV | 6771 | 1154 (17%) | 5617 (83%) |  | 1840 (27%) | 4931 (73%) |  | 2282 (34%) | 4489 (66%) |  |
| Total | 6856 | 1187 | 5669 |  | 1879 | 4977 |  | 2332 | 4524 |  |
| Susceptibility CNVs | 194 | 45 (23%) | 149 (77%) | 1.47* | 69 (36%) | 125 (64%) | 1.48** | 80 (41%) | 114 (59%) | 1.38* |
| No significant or susceptibility CNV | 6771 | 1154 (17%) | 5617 (83%) |  | 1840 (27%) | 4931 (73%) |  | 2282 (34%) | 4489 (66%) |  |
| Total | 6965 | 1199 | 5766 |  | 1909 | 5056 |  | 2362 | 4603 |  |
| Significant or Susceptibility CNVs | 279 | 78 (28%) | 201 (72%) | 1.89**** | 108 (39%) | 171 (61%) | 1.69**** | 130 (47%) | 149 (53%) | 1.72**** |
| No significant or susceptibility CNV | 6771 | 1154 (17%) | 5617 (83%) |  | 1840 (27%) | 4931 (73%) |  | 2282 (34%) | 4489 (66%) |  |
| Total | 7050 | 1232 | 5818 |  | 1948 | 5102 |  | 2412 | 4638 |  |

\*p<0.05; \*\*p<0.01; \*\*\*p<0.001; \*\*\*\*p<0.00001. OR (Odds Ratios) using chi-square test for independence, in relation to not having significant or susceptibility CNV. (%): Row percentages.
<sup>a</sup>High Traits: top 10% gender adjusted t-scores for SWAN (ADHD), TOCS(OCD), CBCL (Anxiety, asked in OSC1), RCADS (Anxiety, asked in OSC2), and/or AQ (ASD, only asked in OSC2).
People with reported diagnoses were not excluded
<sup>b</sup>Reported Diagnosis (DX): ADHD, OCD, ASD, Anxiety, Tics/Tourette's, Learning Problem (OSC1)/Disability(OSC2), Eating Disorder, Mood Disorder, Schizophrenia/Bipolar/MD, Motor Disability(OSC1,OSC2Summer), Velocardiofacial Syndrome, Intellectual Disability (OSC1 & OSC2)/Developmental Delay (OSC1), Language Disorder (OSC1,OSC2 Summer)

Participants carrying either a significant or susceptibility CNV were 2.8 to 1.7 times more likely to report a mental health disorder or have high trait scores (Table 3). Mental health disorders were more common among individuals carrying clinically significant CNVs (*p*=1.9×10^−6^, odds ratio: 3.09 (CI:1.92-4.89)), susceptibility loci (*p*=0.03, odds ratio: 1.47 (CI:1.02-2.08)) or either type of CNVs (*p*=8.46×10^−6^, odds ratio: 1.89 (CI:1.42-2.48)) compared with those did not have any clinically relevant CNVs (Table 3). We observed a similar trend in participants with different types of CNVs with respect to high traits and disorders and high traits combined (Table 3).

We checked to see if participants carrying clinically significant or susceptibility CNVs differed in ADHD, OCD, SSRT or RTV trait scores from those not carrying clinically relevant CNVs (Fig. 1B) using a *t*-test. Comparisons were not statistically significant for OCD or RTV (*p*>0.5). However, participant with susceptibility CNVs had higher ADHD trait scores than those without clinically relevant CNVs (*p*=0.005). Participants with clinically significant CNVs had greater inattention scores than those who did not have any clinically relevant CNVs (*p*=0.006). Participants with susceptibility loci had higher inattention (*p*=0.02) and hyperactivity (*p*=0.005) scores than those without clinically relevant CNVs. Similarly, participants with clinically significant CNVs had higher SSRT scores (poorer inhibitory control) compared to those with susceptibility CNVs (*p*=0.01) or without clinically relevant CNVs (*p*=0.03).

## Clinically relevant CNVs confer risk for a combination of various phenotypes

We presented all clinically significant CNVs and associated phenotypes in Fig. 1C and all susceptibility CNVs in Fig. 1D and 1E (see Supplementary Table S3 for details). Variable expressivity and pleiotropy of a CNV was implied by the observation of different phenotypes associated with the respective CNV. We sub-grouped clinically significant CNVs into aneuploidies, recurrent genomic syndromes and non-recurrent CNVs (*1, 7*). We found aneuploidies in 0.1% (n=7) of European participants but none in East Asian: one female trisomy 21, one triple X syndrome (47,XXX), four XYY syndromes, and one female 45,XO participant. Individuals with XYY syndrome showed a spectrum of different disorders and traits, notably ADHD, ASD, OCD, anxiety, and learning problems, whereas the triple X syndrome participants did not report any NDD/NP phenotype though they reported Celiac Disease (Fig. 1C).

Recurrent CNVs (i.e., the same rearrangements that arise independently in the population) were found in 0.7% (48/7,100) of participants (Table 2 and Supplementary Table S3; Fig. 1C). Examples of these CNVs that were associated with a spectrum of different phenotypes are 15q11-q13 duplication, associated with ASD, OCD, learning problem, and anxiety, 22q11.2 deletion, associated with ASD, ADHD, and learning problem and 16p12.1 distal duplication syndrome with anxiety disorder (Fig. 1C). CNVs impacting *NRXN1* (with ADHD, OCD, anxiety traits, ASD, executive function, and learning problems), *ASTN2* in male (with ADHD, anxiety, and seizure), and *CHD2* (ADHD, learning problems, epilepsy, and hearing problems) were examples of non-recurrent clinically significant CNVs associated with phenotypes (Fig. 1C; Table 2; Supplementary Table S3). Terminal 4q35 deletion, Xq22.3-q23 deletion and deletions impacting *PCDH15*, and *KCNQ5* from this class, did not show any phenotype in our cohort.

Examples of susceptibility CNVs were deletions and duplications impacting *MACROD2 (8)* (with ADHD, anxiety, learning problem, response inhibition and reaction time variability phenotype and family history of mental health condition), *DLGAP2* (with ADHD, anxiety, reaction time phenotype, and family history of mental health condition), *DLG2* (reaction time phenotype), *DPP6* (with anxiety traits), *GRIK2* (with ADHD, reaction time and response inhibition phenotypes and family history of mental health condition), and *CHAMP1* (with ADHD, anxiety, learning problems, and family history of mental health condition) (Fig 1C; Table 2; Supplementary Table S3). Of the latter class, the following CNVs were examples that did not manifest any phenotype: deletions or duplications on *CNTNAP2, ASXL3, ANK2, MBD5, DMXL2*, and *TRIO*. However, the participant with the CNV on *MBD5* duplication is reported as being gifted, and attends a special classroom.

We found 35 unrelated participants carrying duplications impacting either *CNTN6, CNTN4*, or both genes. Sixteen of these participants reported a mental health condition or high traits (Supplementary Tables S3). We performed whole genome sequencing of nineteen samples to search for variants associated with neurodevelopmental disorders but which might have been missed by array (see Supplementary Information), but none were found.

We identified eight participants carrying multiple clinically relevant CNVs (Supplementary Table S3). One male participant of East Asian ancestry had a 28.9 kb duplication overlapping *DLG2* and a 90.3 kb duplication overlapping *TRIP12*, who had slowed reaction time. A female participant of European descent had two recurrent genomic disorder CNVs: a 16p11.2 proximal duplication (604 kb), and a 1q21.1 distal duplication (1.05Mb). She had a family history of ADHD. We also analyzed for clinically relevant SNVs but nothing compelling was found (Supplementary Table S3).

## Replicating our CNV findings in ALSPAC cohort

We replicated clinically relevant CNV findings in an independent pediatric cohort of 7,219 participant (7-17 years of age) of the Avon Longitudinal Study of Parents and Children (ALSPAC; see Supplementary Text; 3,990 European and 3,329 non-European)(*9-11*). ALSPAC is a longitudinal birth cohort, where the participants had been followed to adulthood.

We found NDD/NP clinically relevant CNVs in 3.5% (250/7,219) of ALSPAC participants ranging from 3.2% in European to 3.8% in non-European participants (Supplementary Table S4). The distribution of significant CNVs, susceptibility CNVs, and the combined clinically relevant CNVs among European and non-European are similar to that of the Spit for Science samples (Supplementary Table S4). A previous study found 1.25% of participants carried a clinically relevant CNV in ALSPAC (*12*), but they only analyzed a subset of clinically relevant variants.

## Genetic burden analysis of CNVs

To investigate the relationship of rare CNV variants with mental health traits more generally in Spit for Science, we analyzed rare (<0.5% frequency) and less-rare (1-5% frequency) variants burden using linear regression for number of genes impacted by deletions and duplications, with each trait as the outcome variable and correcting for population stratification, genotyping batch, and genotyping platforms (see Supplementary Methods). Common CNVs (frequency > 5%) were not included in the analysis due to the limited number of variants. For SSRT and RTV, we also accounted for concurrent stimulant medication treatment. European and East Asian participants were analyzed separately due to unbalanced participants ratio (European/Asian=4.0). Trait measurements in Spit for Science and ALSPAC were not comparable. Thus, no further burden analysis was performed on the latter cohort. The number of genes impacted by rare deletions were positively correlated with inattention traits in the European participants (*β*=0.15, 95% CI= [0.01, 0.3], *p*=0.04). We did not find any association between the rare or less rare CNVs and other traits in Europeans or East Asians. While previous studies found an enrichment of deletions in ADHD (*13, 14*), we found a non-significant trend for this association (*β*=0.12, 95% CI= [-0.03, 0.26], *p*=0.12). While a relationship between ADHD and low IQ and between low IQ and large CNVs has been previously described (*15*), low IQ is not likely driving the increase in large CNVs for ADHD in our study. Although we did not measure IQ, there were only two cases with reported intellectual or developmental delay and thus were not likely strong contributors to the findings. Consistent with previous findings with OCD patients, we did not observe association with duplications or deletions (*16*)(Supplementary Table S5).

## Gene-set (pathway) burden analysis of CNVs

We performed burden analysis of rare and less-rare CNVs on 34 gene-sets, representing human neural function and phenotype (n=17), human brain gene/protein expression (n=7), orthologs of mouse genes implicated in nervous system and behavioral phenotypes (n =3) or other organ system phenotypes (n=7); the latter can be treated as putative negative controls (Supplementary Table S5). Gene-set burden was adjudicated using only gene counts per participant and was corrected for global burden (total gene count impacted by deletions and duplications) to avoid nonspecific results, following established practices (*17*).

In the Europeans, we found an increased burden of genes impacted by rare deletions that are highly expressed in brain (*p*=0.02, BH-FDR=0.15), or are in synaptic pathway (*p*=0.03, BH-FDR=0.15), mice neuronal behavior (*p*=0.03, BH-FDR=0.15) and mice nervous system (*p*=0.05, BH-FDR=0.20) in ADHD (Fig. 2; Supplementary Fig. S4; Supplementary Table S5). A similar association was observed with deletions in inattention traits. In East Asians, we found a significant association between the burden of rare duplications impacting genes related to higher mental function and SSRT (*p*=5 × 10^−3^, BH-FDR=0.11). ADHD and hyperactivity were associated with synaptic or neurofunctional genes in the analysis of common deletions (*p*<0.05, BH-FDR<0.2).

**Fig. 2:**
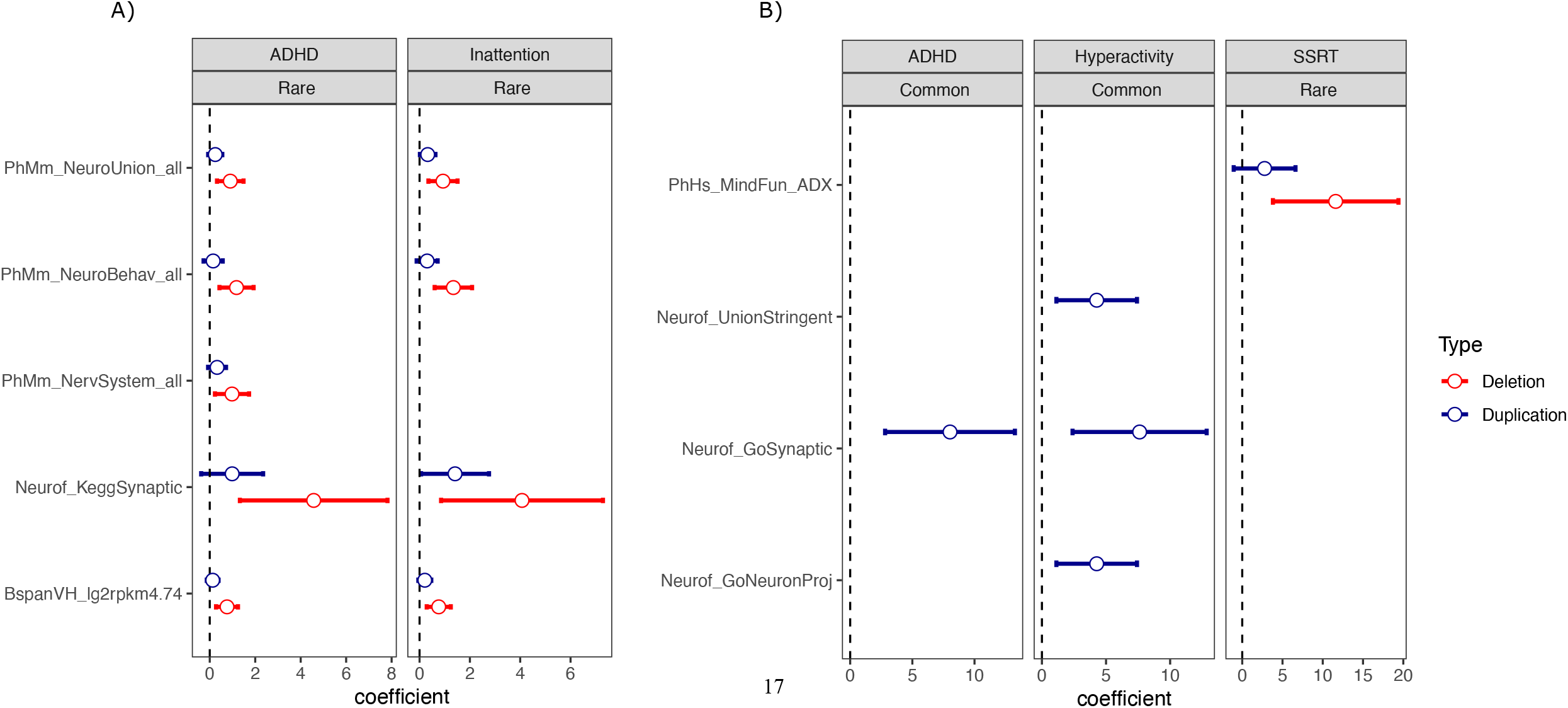
Associated gene-sets identified by gene-set burden analysis of rare (< 0.05 frequency) and common (1.0-5.0 frequency) CNVs separately for deletions and duplications in A) European and East Asian participants The dot represents the beta coefficient from the regression model, and the arm represent 95% confidence intervals of the coefficient. See Supplementary Table S5 for details on gene-sets.

Burden analysis of genes within chromosome X was also performed for individuals of European ancestry. We found a significant association between rare deletion impacting genes that are highly expressed in brain for ADHD (*p*=0.02, BH-FDR=0.11), inattention (*p*=0.04, BH-FDR=0.18), and hyperactivity (*p*=0.04, BH-FDR=0.18).

## Locus burden tests and genes driving signals in gene-set burden analysis

To identify specific rare CNVs associated with mental health disorders or traits, we performed a locus association test on the genes within the top associated gene-sets for each trait (*p*<0.05, PH-FDR<0.2). We present genes with deletions or duplications found in at least 3 individuals and *p*<0.05 for each trait in Table S5. No loci passed genome-wide significant threshold (Family-wiser error rate - adjusted *p* < 0.05). In European subset, we found that rare deletions impacting *ADCYAP1, BBOX1, CSMD1, ZNF74, DGCR6, BCR*, and *GRM6* were associated with ADHD. Similarly, inattention traits were associated with *BBOX1, CSMD1*, and *GRM6* for deletions and 22q11.21 for both deletions and duplications. For the analysis of X-linked genes, we also found rare deletions impacting Xp22.31 region (*PNPLA4* and *STS*) were associated with ADHD, inattention, and hyperactivity-impulsivity. For East Asian subset, we found that 15q11.2 duplication (*CYFIP1*) was significantly associated with ADHD and hyperactivity-impulsivity.

## Conclusions

Our findings demonstrate the value of a community-based sample to help to clarify the role of genetic variation in mental illness. In our approach, we tested the relationship between rare CNV burden and a wide array of traits relevant to NDD/NP including a performance-based cognitive task. We also simplified our reporting of the CNV and clinical impact data in Figure 1, which helps to bring forward the most relevant genomic regions for discussion.

First, we showed that CNVs that are pathogenic based on clinical studies often, but not always, co-occur with notable mental health phenotypes. There are several reasons why there might be no phenotype in individuals with a putative pathological CNV. For example, participants might be too young to manifest adult-onset disorders, failure to report for personal reasons in the context of a community study, or they have CNVs with incomplete penetrance (*3, 7, 18-20*). Although we were not able to clinically verify diagnoses, the prevalence of reported disorders (e.g., ADHD 6.7%) in our sample were similar with existing population figures (*21, 22*). The overall prevalence of diagnoses of mental illnesses in our Spit for Science cohort, i.e. 17.5% in Table 1, was also in accordance with that in the Sweden study (20.9%)(*23*) but less than that (36.7%) in the North Carolina study (*24*). The Sweden study estimate did not include anxiety and depression, which we did account for.

Second, we found clinically significant CNVs with various manifestation of mental health traits/illnesses mainly ADHD, OCD, ASD, and impaired response inhibition. These findings add to mounting evidence that traits share genetic risk with their respective disorders (*25*). Our approach also identified novel variants for each disorder (Supplementary Table S5, showing the added value of the trait-based approach. Another benefit of this approach is examining cognitive deficits present across many psychiatric disorders. Interestingly, response inhibition, which is a common cognitive deficit in ADHD and shares genetic risk with the disorder (*5, 6*), showed overlap in burden findings and specific loci. Many of the CNVs associated with ADHD, OCD and response inhibition traits have been previously implicated in other NDD/NP studies (*4, 26*). Our new findings support evidence from previous studies of common and rare variants of pleiotropy among these disorders (*27-29*).

The current study observed a distribution of clinically relevant CNVs much like those reported in previous child and adult population studies., i.e. ALSPAC (*12*), The Child and Adolescent Twin Study in Sweden (*23*), and The Norwegian Mother, Father, and Child study (*30*). Other studies focused on adults, i.e. UK Biobank (*18, 31*), Estonian Genome Centre at the University of Tartu (*20*), Minnesota Centre for Twin and Family Research (*20*), and Geisinger MyCode community health initiative (*32*) or children and adults (BioMe Biobank)(*33*) has been previously published. However, all these studies focused on recurrent and/or non-recurrent contiguous genomic syndromes only. Several studies examined CNVs in two well-known pathogenic, genes linked to, i.e., *NRXN1* and *SHANK3*, (summary of afore-mentioned studies in Supplementary Table S6)(*12, 18*). Our study was unique in that it evaluated the clinical impact of every CNV including those impacting genes associated with various neuropsychiatric and mental health conditions, which leads us to find all possible clinically relevant CNVs. We explored variable expressivity, pleiotropy (*34*) and the degree of penetrance of each variant because we measured broad and comprehensive phenotypes for participants compared to aforementioned studies. Our phenotype information encompassed various child mental health disorders and medical conditions including diabetes, scoliosis, and ulcerative colitis.

As we have contemplated, somewhat uniquely amongst the medical fields, diagnosis in psychiatry and mental health depends largely on descriptive signs and symptoms, rather than the use of biomarkers (*35*). Here we show specific CNV impacting important neurodevelopmental genes are found in ∼4% of individuals in our community-based sample. From our other research, we anticipate that the clinical findings will nearly double when genome sequencing is used since all classes of genetic variants (sequence-level mutations, smaller CNVs, structural variations, and mitochondrial) are found (*36*). Moreover, including polygenic risk score analysis of common genetic variants (*37-39*) or gene conservation weighting (*26, 40*) may help to further delineate phenotypic expression of traits in penetrant CNV carriers. With the number of people who receive a clinical diagnosis of a mental illness on the rise and new CNVs/genes constantly being implicated in these conditions, irrespective of possible confounding variable expressivity and penetrance, we believe there is value to actively integrate the genomic etiological information (Figure 1) into the clinical diagnosis paradigm.

## Data Availability

The relevant genotype data and CNVs are being prepared for submission to the European Genome-Phenome Archive and the dbVar Database, respectively.

## Acknowledgements

The authors thank The Centre for Applied Genomics at The Hospital for Sick Children for technical assistance. We also thank the families who participated in ALSPAC study, and the ALSPAC team. This study was funded by operating grants from the Canadian Institutes of Health Research award to PDA (MOP-106573) and RJS (MOP–93696). It was also supported by grants from Genome Canada, Canada Foundation for Innovation, Government of Ontario, Canadian Institutes of Health Research, The Hospital for Sick Children Foundation and the University of Toronto McLaughlin Centre. The UK Medical Research Council and Wellcome (Grant ref: 217065/Z/19/Z) and the University of Bristol provide core support for ALSPAC. RJS holds the Toronto Dominion Bank Financial Group Chair in Child and Adolescent Psychiatry. PDA holds the Alberta Innovates Health Solutions (AIHS) Translational Health Chair in Child and Youth Mental Health at the University of Calgary. SWS holds the Northbridge Chair in Paediatric Research, The Hospital for Sick Children and the University of Toronto.

## Conflict of Interest Statement

RJS consults for Highland Therapeutics and Purdue Pharma. SWS is on the Scientific Advisory Committees of Population Bio, and serves as an Academic Consultant for the King Abdulaziz University. These relationships did not influence data interpretation or presentation during this study. The other authors declare no conflict of interest.

## Ethics Approval

The Research Ethics Board of The Hospital for Sick Children gave ethical approval for this work. Informed consent, and verbal assent where applicable, was obtained from all participants and/or their legal representatives to take part in this study and to have this research work published.

## Notes

### Funding Statement

This study was funded by operating grants from the Canadian Institutes of Health Research award to PDA (MOP106573) and RJS (MOP93696). It was also supported by grants from Genome Canada, Canada Foundation for Innovation, Government of Ontario, Canadian Institutes of Health Research, The Hospital for Sick Children Foundation and the University of Toronto McLaughlin Centre. The UK Medical Research Council and Wellcome (Grant ref: 217065/Z/19/Z) and the University of Bristol provide core support for ALSPAC. RJS holds the Toronto Dominion Bank Financial Group Chair in Child and Adolescent Psychiatry. PDA holds the Alberta Innovates Health Solutions (AIHS) Translational Health Chair in Child and Youth Mental Health at the University of Calgary. SWS holds the Northbridge Chair in Paediatric Research, The Hospital for Sick Children and the University of Toronto.

### Author Declarations

The Research Ethics Board of The Hospital for Sick Children gave ethical approval for this work.

