## Supplemental Materials for "Gene copy number variation in pediatric mental illness in a general population"

**Short titles:** An exploration of CNVs in a diverse pediatric cohort

Mehdi Zarrei[1,2,†], Christie L. Burton[3,†], Worrawat Engchuan[1,2,†], Edward J Higginbotham[1,2], John Wei[1], Sabah Shaikh[3], Nicole M. Roslin[1,2,3], Jeffrey R MacDonald[1], Giovanna Pellecchia[1], Thomas Nalpathamkalam[1], Sylvia Lamoureux[1], Roozbeh Manshaei[1,4], Jennifer Howe[1], Brett Trost[1,2], Bhooma Thiruvahindrapuram[1], Christian R Marshall[1,5,6], Ryan KC Yuen[1,2], Richard F. Wintle[1], Lisa J Strug[1,2,7], Dimitri J. Stavropoulos[5], Jacob A.S. Vorstman[8,9], Paul Arnold[2,10,11], Daniele Merico[1,12], Marc Woodbury-Smith[1,13], Jennifer Crosbie[3,8,‡], Russell Schachar[3,8,14,‡], Stephen W. Scherer[1,2,15, ‡,*]

[1]The Centre for Applied Genomics, The Hospital for Sick Children, Toronto, ON, Canada.
[2]Program in Genetics and Genome Biology, The Hospital for Sick Children, Toronto, ON, Canada.
[3]Neurosciences and Mental Health Program, The Hospital for Sick Children, Toronto, ON, Canada.
[4]Ted Rogers Centre for Heart Research, Cardiac Genome Clinic, The Hospital for Sick Children, Toronto, Ontario, Canada.
[5]Genome Diagnostics, Department of Paediatric Laboratory Medicine, The Hospital for Sick Children, Toronto, ON, Canada.
[6]Laboratory Medicine and Pathobiology, University of Toronto, Toronto, ON, Canada.
[7]Division of Biostatistics, Dalla Lana School of Public Health, University of Toronto, Toronto, Canada.
[8]Department of Psychiatry, University of Toronto, Toronto, ON, Canada.
[9]Autism Research Unit, The Hospital for Sick Children, Toronto, ON, Canada.
[10]Mathison Centre for Mental Health Research and Education, University of Calgary, Calgary, AB, Canada.
[11]Departments of Psychiatry & Medical Genetics, Hotchkiss Brain Institute, Cumming School of Medicine, University of Calgary, Calgary, AB, Canada.
[12]Deep Genomics Inc., Toronto, ON, Canada.
[13]Institute of Neuroscience, Newcastle University, Newcastle upon Tyne, UK.
[14]Institute of Medical Science, University of Toronto, Toronto, ON, Canada.
[15]Department of Molecular Genetics and McLaughlin Centre, University of Toronto, Toronto, ON, Canada.
[†]These authors contributed equally
[‡]These authors share senior authorship

### Materials and Methods

#### *Participants*

Spit for Science is a community-based study of 22,515 children and adolescents 4-18 years of age (mean age 10.7, SD=2.9, 50.3% male) seen at the local Science Centre (OSC) from 2008 to 2019 and was then paused due to the pandemic. There were two phases of data collection (a) Spit for Science 1 (OSC1), collected between July 2008 and September 2009 (n=17,264), and (b) Spit for Science 2 (OSC2), collected between June 2019 and March 2020 (n=5,743).

Informed consent, and verbal assent where applicable, approved by The Hospital for Sick Children's Research Ethics Board was obtained from all participants. Parents provided information about community diagnosis and history, and rated child behavior (see below) for 81.8% of participants. Older, unaccompanied youth provided information about themselves (16.9%) if they were capable of consenting.

#### *Behavioral measures*

Using a computerized questionnaire, participants reported demographic, medical, mental health, educational and family history information (*6*). We also asked if participants had ever spent time in a special classroom (gifted, special education, resource room, learning disability, language impairment, ADHD, or behavioral issues) and if they had ever been diagnosed or been treated for a learning problem. See below for details about each measure. This is the question we asked:

Do [nickname]'s biological Parent(s) or Sibling(s), who are not participating in the study, have a history of mental health issues (i.e. diagnosis of or treatment for)? ex. anxiety, learning problems, etc. If Yes, please specify below:

#### *Mental health disorder*

Participants were considered to have a mental health / neurodevelopmental disorder if they or their parent reported that they received a diagnosis of or treatment for attention-deficit/hyperactivity disorder (ADHD), obsessive-compulsive disorder (OCD), autism spectrum disorder (ASD), Tourette's, or any other neurodevelopmental or neuropsychiatric disorder (e.g., intellectual disability, learning problems, developmental delay, cerebral palsy, mood and anxiety disorders).

#### *Mental health trait measures*

<u>ADHD Traits</u>

The Strengths and Weaknesses of ADHD Symptoms and Normal Behavior Rating (SWAN) Scale (*8, 9*) was used to measure ADHD traits (inattention and hyperactivity-impulsivity). The SWAN is based on the 18 ADHD items from the Diagnostic and Statistical Manual of Mental Disorders, 4$^{th}$ edition (DSM-IV)(*1*) and items are worded to permit informants to report both "strengths" and "weaknesses" on a seven-point scale (*2*). For example, -3 = far above average indicates low ADHD trait scores to +3 = far below average indicates the presence of an ADHD trait. Values of 0 indicate average levels of each item. The measure generates a total score (-54 to +54) and two subscales for inattentive and hyperactive/impulsive traits (-27 to +27 for each sub-scale). The SWAN has high internal consistency, test-retest reliability, as well as convergent, divergent and discriminant validity (*3*).

A participant was considered to have 'High ADHD traits' if they fell within the top 10% of the t-scores for SWAN total score, SWAN Hyperactive/Impulsive, and/or SWAN Inattentive scales. Participants with community-reported diagnosis of ADHD were included in 'High ADHD traits" if their t-scores fell within the top 10% of either SWAN Total, SWAN-HI, and/or SWAN IA scales.

<u>OCD Traits</u>

Traits relevant to OCD was measured with the Toronto Obsessive-Compulsive Scale (TOCS) (*7*). TOCS includes 21 items scored on a scale of -3 to +3 (-3 far less often than average; -2 less often than average; -1 slightly less often than average; 0 average amount of time; 1 slightly more often than average; 2 more often than average; and 3 far more often than average). The TOCS has excellent internal consistency, inter-rated reliability, and divergent and convergent validity (*4*).

A participant was considered to have 'High OCD traits' if they fell within the top 10% of the t-scores for TOCS total score. Participants could have a community-reported diagnosis of OCD and still be considered to have 'High OCD Traits' if their t-scores fell within the top 10% of TOCS total score.

<u>Anxiety Traits</u>

For Spit for Science 1, we measured anxiety traits using the Child Behavior Checklist (CBCL) anxiety problems sub-scale which has 9 items based on DSM-5 that each range from 0 ('not true') to 2 ('very true or often true') now or in the past 6 months.

For Spit for Science 2, we measured anxiety traits using the generalized anxiety sub-scale Revised Children's Anxiety and Depression Scale (RCADS-25; Ebesutani et al., 2012;2017) comprising of 15 anxiety items. Items were rated on a 4-point Likert scale (1 = never; 4 = always). Standardized t-scores controlling for gender , grade, and respondent (parent vs self) were calculated based on the total anxiety scores.

A participant was considered to have "High Anxiety Traits" if they scored within the top 10% of t-scores of the grouped data.

<u>ASD Traits</u>

ASD traits were not measures in Spit for Science 1. In Spit for Science 2, we measured ASD traits using and abridged version of the Autism Spectrum Quotient-28 (AQ-28)(*5*) selecting the same items as those from the AQ-short for adults. Items were based on a 4-point Likert scale ranging from 0-3, indicating high and low trait levels, with four response options ranging from "strongly disagree" to "strongly agree". Items were grouped using the five factors identified by Hoekstra et al. (*6*): Switching, Social Skills, Routine, Numbers and Patterns, and Imagination. However, since Numbers and Patterns did not correlate without other factors, items in this category were dropped in this analysis (n=5).

A participant was said to have "High ASD traits" if they fell withing the top 10% of AQ total t-scores.

<u>Stop signal task</u>

Participants performed a twelve-minute computerized stop-signal task (SST) to measure response inhibition (stop-signal reaction time) and reaction time variability (RTV) (*7*) described in detail elsewhere (*8*). Response inhibition and reaction time variability have been associated with ADHD and OCD, are heritable traits, and share genetic risk with ADHD (*10*)(*6*). Longer stop-signal reaction time (SSRT) reflects poorer response inhibition and greater RTV reflects greater moment-by-moment fluctuation in attention.

The SST involves two types of trials, go and stop trials. On go trials, subjects were instructed to respond using a left-hand (X) or right-hand (O) key as quickly and as accurately as possible to X's and O's presented on a computer screen one at a time using a video game controller. Stop trials involve presentation of a 1,000 Hz tone through headphones on a random subset of 25% of go trials. Participants were instructed to stop their response when they heard the stop signal if they could. The initial delay between the presentation of go stimuli and stop signals was set at 250 ms. The length of this delay was dynamically adjusted depending on participant performance; the delay was shortened by 50ms if the participant was unable to stop or lengthened by 50 ms if they were able to stop. There was a practice block of trials (24 trials; 18 go trials and 6 stop trials) and 4 blocks of 24 experimental trials for a total of 72 go trials and 24 stop trials.

We calculated the latency of response inhibition (stop-signal reaction time: SSRT) using interpolation (Verbruggen et al., 2019) and standard deviation of response times (RTV) across all go trials following a correct go response. We considered the distribution of RTV after correct-go responses only and not after all trial types because of skewing due to slowed responses after stop-inhibit (successful stopping) and stop-respond (unsuccessful stopping) trials (Dupuis et al., 2018). Those longer reaction times would confound and inflate RTV by adding between-trial type differences to the estimate of overall variability.

Participants were considered to have poor response inhibition or variable reaction time if they fell within the top 10% of t-scores in any of these categories.

*Standardized Scores*

We created standardized t-scores for SWAN total score, inattentive total score, hyperactive/impulsive total score, TOCS total score, CBCL Anxiety Problems (Spit for Science 1), RCADS Anxiety (Spit for Science 2), AQ (Spit for Science 2), SSRT and RTV. Total scores were modelled using linear regression controlling for age and gender, for parent- and self-respondents separately and residual scores were obtained. Participants missing gender, or self-respondents under 12 years of age were removed from the sample. The final scores had a mean of 50 and a standard deviation of 50.

**Preparation of Genetic Samples and Genotyping**

For Spit 1, DNA was extracted manually from saliva using the Oragene Reagent system and protocol (DNA Genotek, Ottawa, Canada). To precipitate any possible carbohydrates in the sample, we centrifuged the samples for an additional 10 min at 10,000 RPM and removed any formed pellet from the sample. DNA was quantified using the Quanti-iT, Pico Green® dsDNA kit from Invitrogen (Thermo Fisher Scientific, Waltham, MA, USA) and samples with concentrations <60ng/µl were excluded (6.5% of all extracted cases). DNA was subsequently aliquoted and stored at -80°C. Prior to conducting the microarrays, DNA quality was verified using agarose gels and 98.5% of samples had sufficient DNA quality. Samples were genotyped on the Illumina HumanCoreExome-12 BeadChip (v1-0) (Illumina, San Diego, CA, USA).

For Spit 2, DNA was extracted using the chemagic STAR robot using standard Oragene protocols. We used a Biotek FLx800 Multi Detection Microplate Fluorometer to measure the dsDNA concentration with Hoechst dye and samples with concentrations <30ng/ul were excluded (0.95% of all extracted cases). DNA was subsequently aliquoted and stored at -80°C. Samples were genotyped on the Illumina Global Screening Array

All samples in this study were genotyped at The Centre for Applied Genomics (The Hospital for Sick Children, Toronto, ON, CA). In addition, on each 96 well plate we genotyped an individual from a European ancestry HapMap trio as an additional step of quality assurance (*2*). Genotypes were called separately for the HumanCoreExome (GenomeStudio v1.9.4) and Global Screening arrays (GenomeStudio v2.0).

***Genotyping QC***

All SNP position information was based on the GRCh37 reference. Samples were excluded if their call rate was <97%, a heterozygosity rate >6 times the interquartile range from the closest quartile and/or their predicted and reported sex mismatched. SNPs were excluded if they had call rates <97%, the observed genotypes deviated from the proportions expected under the rules of Hardy-Weinberg equilibrium at a false discovery rate (FDR) <1% (using samples homogeneous for ancestry), and/or were duplicates of other SNPs on the array, based on physical position and alleles (the SNP with the highest call rate was retained). Concordance of the HapMap control samples was >99.99%.

Since there are few overlapping SNPs between the HumanCoreExome and Global Screening arrays, both datasets were imputed to the 1000 Genomes reference panel (phase 3 version 5a) using BEAGLE 4.1 (*9*) separately by genotype array and ethnic group. Variants were excluded if they had alleles ambiguous for strand information, were insertion/deletions, had imputation quality score $r^2<0.8$ or minor allele frequency <0.1 in any of the subsets. Dosages were converted to hard calls.

Individuals were assigned to an ethnic background based on the self-reported ancestry of all four grandparents. In order to detect individuals whose inferred ancestry did not agree with their self-report, or whose recent ancestry appeared to come from multiple continents, ancestry proportions were estimated using Admixture 1.3.0. A supervised analysis was performed, using cleaned 1000 Genomes Omni 2.1 array genotypes as a reference, and 6 ancestral populations (AFR, EAS, SAS, AMR, NFE and FIN). Prior to the Admixture analysis, the number of variants was reduced to those present in both datasets (matching on chromosome, physical position and alleles), on the autosomes, not in the major histocompatibility (MHC) region on chromosome 6 or in the region of a known polymorphic inversion on chromosome 8, minor allele frequency >1% (in all data combined), and pairwise linkage disequilibrium $r^2$ <0.4 on a chromosome, leaving 108,321 SNPs. Individuals were removed if they appeared to have mixed ancestry (maximum fraction <0.6) or if their estimated ancestry differed from their self-report.

In order to detect closely related individuals, pairwise kinship coefficients were estimated using KING 2.2.6 (using the --related function). Identical pairs that were not known to be twins based on self-report were removed; for pairs that reported being twins, one individual was selected randomly from the pair. Among the remaining families, 1,263 individuals clustered into 599 family structures containing 1st and 2nd degree relatives. One individual from each family was selected at random, and the rest were discarded. There was a total of 7,100 people used in analyses: 5,686 of European ancestry (4359 from Spit 1 and 553 from Spit 2) and 1,414 of East Asian ancestry (861 from Spit 1 and 553 from Spit 2).

Principal components were estimated separately within each ancestral group using Eigensoft 6.0. For both EUR and EAS, the Spit 1 and Spit 2 samples appeared to be well-mixed.

***(10)Genotyping, CNV calling, and detection of rare variants***

***Quality control filtering***

We genotyped 5,688 participants (Supplementary Table S1) on the Illumina Infinium® HumanCoreExome-12 BeadChip array (Illumina Inc., San Diego, CA, USA). Quality controls of CNVs were performed as previously described (*11, 12*), and 418 samples failed our stringent quality control criteria. CNVs were called using three algorithms: PennCNV (*13*), QuantiSNP (*14*), and iPattern (*15*). Since PennCNV and QuantiSNP are not able to call CNV on chromosome Y, deletions and duplications on chromosome Y were not included in the statistical analyses. Since QuantiSNP doesn't perform well on chromosome X for this dataset, i.e. under calls or over calls CNVs depending on the sample, we only used chromosome X calls from PennCNV and iPattern. We defined a 'stringent' set of rare CNV, those detected by a minimum of two algorithms (Supplementary Table S1). Since these algorithms are not particularly suitable for calling CNVs > 10Mb (*11, 12, 16*), e.g. duplication of chromosome X in cases with Klinefelter syndrome, these large CNVs can be fragmented into multiple calls. These were merged manually and their identity confirmed by examining the probe density and B allele frequency plots in the region; specifically, neighboring variants from the same sample with the same copy number (deletions or duplications) are merged together if the distance between them was < 50kb. We additionally inspected our data for possible sex chromosome aneuploidies that might have been missed in our regular pipeline. To do this, we analyzed heterozygosity ratio on chromosome X SNPs and call rate on chromosome Y SNPs. For these CNVs, B allele frequency plots were inspected manually for further confirmation of their validity. We focused our analyses on CNVs with a minimum of 10 kb in size and identified by at least five consecutive probes. While CNVs on the Y chromosome were not included in downstream analyses (discussed above), we presented and discussed chromosome Y aneuploidies in the paper. The genomic coordinates used are based on Human Genome Build GRCh37/hg19. We found 21 samples where the genetic data predict the sex as female, but they are reported as male by self- or parental-report (Supplementary Table S1).

To identify the frequency of the CNVs, we used a two-step process. First, the population frequency of CNVs was derived using 10,851 population control samples of different ethnicities (explained in details elsewhere)(*11, 12, 16-18*). Second, we calculated the internal frequency separately for each population group. In the burden analysis, we retained only CNVs with population and internal frequency of less than 5% and not overlapping DGV Gold Standard CNV clusters (*19*). For all these CNV frequency calculations, we used a 50% reciprocal overlap strategy (*20*). We additionally removed CNVs with more than 70% overlap with repeat regions of the genome and low complexity DNA sequences. CNVs in Asian samples were also filtered by an additional population specific control CNVs set. This set included 1,679 Asian subjects compiled from multiple sources: 919 samples from Singapore database (*21*), 101 samples from HapMap project (*22*), 451 samples from Lu et al. (*23*), and 206 parental samples from Gazzellone et al (*17*).

We identified 14,809 rare CNVs defined as having lower than 1% frequency in the dataset (2.06/individual; Supplementary Table S1), of which 7,256 (49 %) were deletions and 7,553 (51 %) were duplications.

### *SNPs calling and detection of rare, prioritized variants*

Genotypes were called using GenomeStudio (Illumina, San Diego, CA, USA). QC for SNPs followed the standard guidelines provided by the software. Briefly, individuals with <0.95 genotype calls were excluded from the downstream analysis. Probes with >5% missingnesses or that deviated from Hardy-Weinberg equilibrium ($p< 1.0\times 10^{-6}$) were removed.

Frequency of SNVs/indels were defined against multiple external controls datasets (1,000 Genomes, gnomAD exomes, and gnomAD genomes) and frequency < 0.1% in this data-set. SNPs overlapping segmental duplications were removed as they may be enriched in genotyping artifacts. SNPs were categorized as LOF (frameshift indels, splicing and stop-gain variants) or missense using Annovar (*24*). A damaging missense variant was defined as passing five out of seven damaging prediction scores cutoffs: SIFT < 0.01, PolyPhen >= 0.90, MutationTaster >= 1.90, PhyloP Mammalian >= 2.30, PhyloP 100 Vertebrates >= 4.0, CADD Phred >= 15, and if variant is on pfam domains or PhastCons elements (see Yuen et al. (*25*) for details)(Supplementary Table S1).

### *Whole genome sequencing*

Nineteen unrelated participants carrying duplications impacting *CNTN6* or *CNTN4* were whole genome sequenced on the Illumina HiSeqX following the manufacturer's recommended protocol (Illumina Inc., San Diego, CA, USA). We first used NEBNext Microbiome DNA Enrichment Kit to separate human DNA from the microbiome DNA, allowing the human DNA to be recovered by the beads. About 1 ug of human-enriched genomic DNA was submitted to The Centre for Applied Genomics for genomic library preparation and whole genome sequencing. DNA samples were quantified using Qubit High Sensitivity Assay and sample purity was checked using Nanodrop OD260/280 ratio. One-hundred ng of DNA was used as input material for library preparation using the Illumina TruSeq Nano DNA Library Prep Kit, following the manufacturer's recommended protocol. In brief, DNA was fragmented to 400 bp on average using sonication on a Covaris LE220 instrument; fragmented DNA were then end-repaired, A-tailed and indexed TruSeq Illumina adapters with overhang-T were added to the DNA. Libraries were validated on a Bioanalyzer DNA High Sensitivity chip to check for size and absence of primer dimers, and quantified by qPCR using Kapa Library Quantification Illumina/ABI Prism Kit protocol (KAPA Biosystems). Validated libraries were pooled in equimolar quantities and paired-end sequenced on an Illumina HiSeqX platform following Illumina's recommended protocol to generate paired-end reads of 150-bases in length.

We detected CNVs from WGS for each samples using two algorithms, CNVnator (*26*) and ERDS (*27*), as previously described (*25, 28*). We retained CNVs ≥ 1kb. We defined a stringent set of CNVs as those called by both CNVnator and ERDS, i.e. with 50% reciprocally overlapped. Rare CNVs were defined as those with <1% frequency in the WGS control CNVs (defined as parents in MSSNG cohort (*25*)), <1% frequency in 10,851 microarray controls (details in (*11*)), overlapping with a region of the genome that is at least 75% copy-number stable according to the CNV map of the human genome (*20*), and are less than 70% overlapped by segmental duplications. They were further refined as those not overlapping with 1% CNV clusters in DGV (*19*)( Supplementary Table S1).

Alignment, variant calling, and detection of SNVs were performed as previously described (*25*). Briefly, reads were aligned to the reference genome (build GRCh37) using Burrows-

Wheeler Aligner (BWA, version v0.7.12). SNVs and indels were detected using the Genome Analysis Toolkit (GATK; version v3.7) HaplotypeCaller. Variants were annotated using a pipeline based on Annovar, as previously described (*25*)(Supplementary Table S1).

Rare SNVs and indels were defined those with <0.1% frequency in the Genome Aggregation Consortium (gnomAD)(*29*) and 1000 Genomes (*30*). LOF and damaging missense variants were identified as explained above.

#### *Interpretation of Clinically Relevant CNVs, and variants associated with known genomic regions*

We analyzed our data for aneuploidies, CNVs at known genomic disorder loci (including DECIPHER syndrome loci (*31*) and ClinGen recurrent regions (*32*)), CNVs greater than 3Mb in size, and pathogenic and risk CNVs associated with a neurodevelopmental and/or mental health phenotypes following the methods described by Zarrei et al.(*11*). To identify pathogenic and risk CNVs, we considered all rare copy number losses that impacted the coding sequence of known ASD-associated and ASD-candidate genes or loci (*25*), and genes associated with an autosomal dominant neurodevelopmental or neuropsychiatric phenotype in the Clinical Genomics Database (*33*) and Online Mendelian Inheritance in Man (OMIM) databases (*34*). CNVs were classified following the guidelines outlined by the American College of Medical Genetics (ACMG)(*35, 36*) (Supplementary Table S2).

#### *Global burden and functional gene-set burden analysis in CNVs*

We performed a global burden and functional gene-set burden analysis for unrelated individuals together for two study phases but separately for European (EUR) and East Asian (EAS) subsets. The test was done using linear regression for three different traits, i.e. ADHD, OCD, and SSRT and RTV. We also analyzed hyperactivity and inattentiveness, two sub-phenotypes of ADHD using the same subjects of the main ADHD trait. Over 88% of subjects (6,164) were shared between the three traits. Only CNVs with frequency <5% and size < 3Mb were included in this analysis. CNVs were annotated with genes whenever they overlapped at least one RefSeq exon of the gene (RefSeq genes (*37*); downloaded August 07, 2019).

Subjects were genotyped in 67 - well plates. When we analyzed the T-score distribution by plate, we found some plates had skewed T-score distributions on ADHD, OCD and stop task variables (Supplementary Figure S2). Consequently, a plate variable was included as a covariate for the burden analysis.

Global burden was analyzed for deletions and duplications separately. CNV burden was quantified using the number of genes impacted per participant. Burden was tested using linear regression, with participant as statistical sampling units for both deletions and duplications. Each trait (ADHD, OCD, stop task SSRT, hyperactivity, inattentiveness and stop task variables SSRT and RTV) was treated as an outcome variable. The first three principal components were included as covariates to correct for population stratification; study phase and genotyping batch were also included as covariates to correct for the potential batch effect previously described (*38*). The burden effect size was estimated as the regression coefficients in model #1; burden significances were derived from Wald test of coefficients.

Model # 1: $y = \alpha C + \beta l L$

Where, y is the phenotypic trait score (T-score) variable; $C = [C1, C2, \ldots, Cn\text{-}1, Cn]$ is the vector of correction covariates (PC1, PC2, PC3, genotyping batch, study phase) and $\alpha$ is a corresponding vector of coefficients; $L$ is a vector of the two main variables (deletions and duplications) to be tested, encoding CNV burden as number of genes impacted by deletions and duplications CNVs, and $\beta l$ are their coefficient. The linear regression was performed using *lm* functions in R (*39*). The 95% confidence interval of the coefficient was estimated using the *confint* R function from the *stats* package.

**Gene-set burden enrichment analysis**

We obtained gene-sets compiled elsewhere (*38*). These included 17 gene-sets representing neural function and phenotypes in human, 5 gene-sets representing brain expression, and 10 gene-sets representing mouse phenotypes in different organ systems including the nervous system. One gene-set, brain specific expression, named here "protein expression" (originally named "blue module"), was compiled in (*40*).

Gene-set burden was tested including a correction for global burden, to ensure the gene-set signal is specific (*41*), Control for global burden was applied to all gene-set burden tests for sake of uniformity but was particularly important for inattention scores because that trait was associated with a significantly greater global burden.

Model # 2: $y = \alpha C + \beta_l L + \beta_s S$

Where, S is the vector with two counts of 1) the genes in the gene-set overlapped by deletions, 2) the number of genes in the gene-set overlapped by duplications. $\beta_s$ is a vector of their corresponding coefficients. The 95% confidence interval of the coefficient was estimated in the same way as described for global burden. Gene-set burden significance was estimated by comparing model #2 and model #1, using the chi-square deviance test, which was done using the anova.lm R function from the stats package v. 3.3.3. We only performed the test on the gene-sets in which at least 3 individuals had CNVs impacting genes in the gene-sets. Multiple test correction by Benjamini Hochberg False Discovery Rate (BH-FDR) was performed using the *p.adjust* R function with *method* = "*BH*". The correction was performed separately for each combination of phenotypic trait and group of gene-sets (neural function, phenotypes and brain expression were corrected separately from mouse phenotypes gene sets). Results of our gene-set burden enrichment analysis are shown in Supplementary Table S4.

***Locus association test***

We conducted locus association tests to identify trait-associated loci and also to find out which genes were driving the signal of significant gene-sets. A gene-level test was first performed, testing the burdens of deletions and duplications for each gene in the significant gene-sets (BH-FDR<20%). For CNVs, genes were then merged into loci whenever they had a highly similar set of rare CNVs (Jaccard's index > 0.35). This procedure has the advantage of grouping highly correlated blocks of genes (which arise in presence of large CNVs) and leads to a more accurate BH-FDR calculation (*38*). Only the genes with at least three individuals had

CNVs overlapping their exon were tested. The gene-level test was performed by fitting a linear regression model with the trait score as outcome variable, the same correction covariates used for the gene-set burden, and the main variable encodes the presence or absence of a rare variant overlapping at least one exon of the gene (in each subject). Significance was estimated comparing model # 3 and model # 1.

Model # 3: $y = \alpha C + \beta_g G$

Where G is two binary variables indicating whether a subject has deletions or duplications overlapping an exon of that gene or not. $\beta_g$ are their corresponding coefficients. We used the locus association results to sort genes within significant gene-sets.

***Investigation of frequency cut-offs for burden analysis***

As the prevalence of traits analyzed are not very rare (e.g. 6.72% ADHD cases in this cohort), we investigated the phenotypic effect of CNVS within four different frequency bins, i.e. (1) 6,232 CNVs with frequency < 0.1%, (2) 3,311 CNVs with frequency between 0.1%-0.5%, (3) 1,593 CNVs with frequency between 0.5%-1% and (4) 2,993 CNVs with frequency between 1%-5%. We did not include CNVs with frequency greater than 5% due to the limited number of CNVs available (698 CNVs). Using the European subset, for each frequency bin, the global burden analysis was done for all traits. The results are shown in Supplementary Figure S3 and Supplementary Table S4. Spearman correlation found that CNVS with frequency <0.1% and 0.1%-0.5% had similar association to T-scores (Spearman's Rho=0.83). We also found a moderate correlation between CNVs with frequency <0.1% and 1%-5% (Spearman's Rho=0.54). Based on this finding, we decided to perform the subsequent analyses for CNVs with frequency <0.5% as rare CNVs and CNVs with frequency 1%-5% as less rare CNVs. We did not include CNVs with frequency between 0.5%-1% in the subsequent analysis due to the limited number of CNVs available.

**Replication sample: Avon Longitudinal Study of Parents and Children (ASLPAC) CNVs**

Pregnant women resident in Avon, UK with expected dates of delivery 1st April 1991 to 31st December 1992 were invited to take part in the study. An initial number of 14,541 pregnant women resident in Avon, UK with expected dates of delivery from April 1991 to December 1992 (*42, 43*). Of these initial pregnancies, there was 14,062 live births and 13,988 children who were alive at 1 year of age (*42, 43*). Samples were genotyped on Illumina Hap550 Quad platform, which is like the platforms used in our study with respect to probe count and distribution. Ethical approval for the study was obtained from the ALSPAC Ethics and Law Committee and the Local Research Ethics Committees. Consent for biological samples has been collected in accordance with the Human Tissue Act (2004). Informed consent for the use of data collected via questionnaires and clinics was obtained from participants following the recommendations of the ALSPAC Ethics and Law Committee at the time.

Copy number variations from 7,590 participants in the ALSPAC cohort were analyzed (*44*) (Supplementary Table S3). The cohort description and the main principles of quality control information are provided in Guyatt et al. (2018)(*44, 45*). Samples were genotyped on Illumina Hap550 Quad platform and the CNVs were called using PennCNV(*13*). We performed quality control on the CNVs calls as outlined above for the Spit for Science cohort. 371 samples did not passed our stringent quality control. Rare CNVs were identified as described for Spit for Science cohort above. Because the CNVs was identified using only one algorithm and we did not have access to DNA samples for validating CNVs, we took a conservative approach to analyze only rare high quality calls, defined as those larger than 50kb and spanning five or more probes. The ASLPAC cohort did not call CNVs on sex chromosomes. We did not perform trait burden analysis in ALSPAC because the measurement of the traits were not consistent between Spit for Science and ALSPAC. Please note that the study website contains details of all the data that is available through a fully searchable data dictionary and variable search tool: http://www.bristol.ac.uk/alspac/researchers/our-data/

***Clinically relevant SNVs***

We searched for clinically relevant LOF mutations and predicted damaging missense mutations on samples genotyped on Core Exome Array. We found two such LOF mutations in two different East Asian participants [a heterozygous canonical splice-site mutation in *EHMT1*: NM_024757:c.1170+2T>C:p.(?) and another heterozygous stop-gain mutation in *TCF12*: NM_207037:c.C1267T:p.(R423X)]. The former participant showed no phenotype whereas the later one had OCD. We also found a heterozygous stop-gain mutation [*GNAS*: NM_080425:c.C538T:p.(Q180X)] in a European participant with poor response inhibition (Supplementary Table S3). These account for 0.05% (3/5,270) of participants. None of the subjects with clinically relevant LOF mutations had clinically relevant CNVs. The three sequence-level variants were confirmed using Sanger sequencing. However, none of clinically relevant variants from Global Screening Array was confirmed by Sanger and therefore were not analyzed. These results are consistent with previous reports of high false positive rates of clinical variants from the (51%) in Global Screening Array (*46*).

**Supplementary Figures**

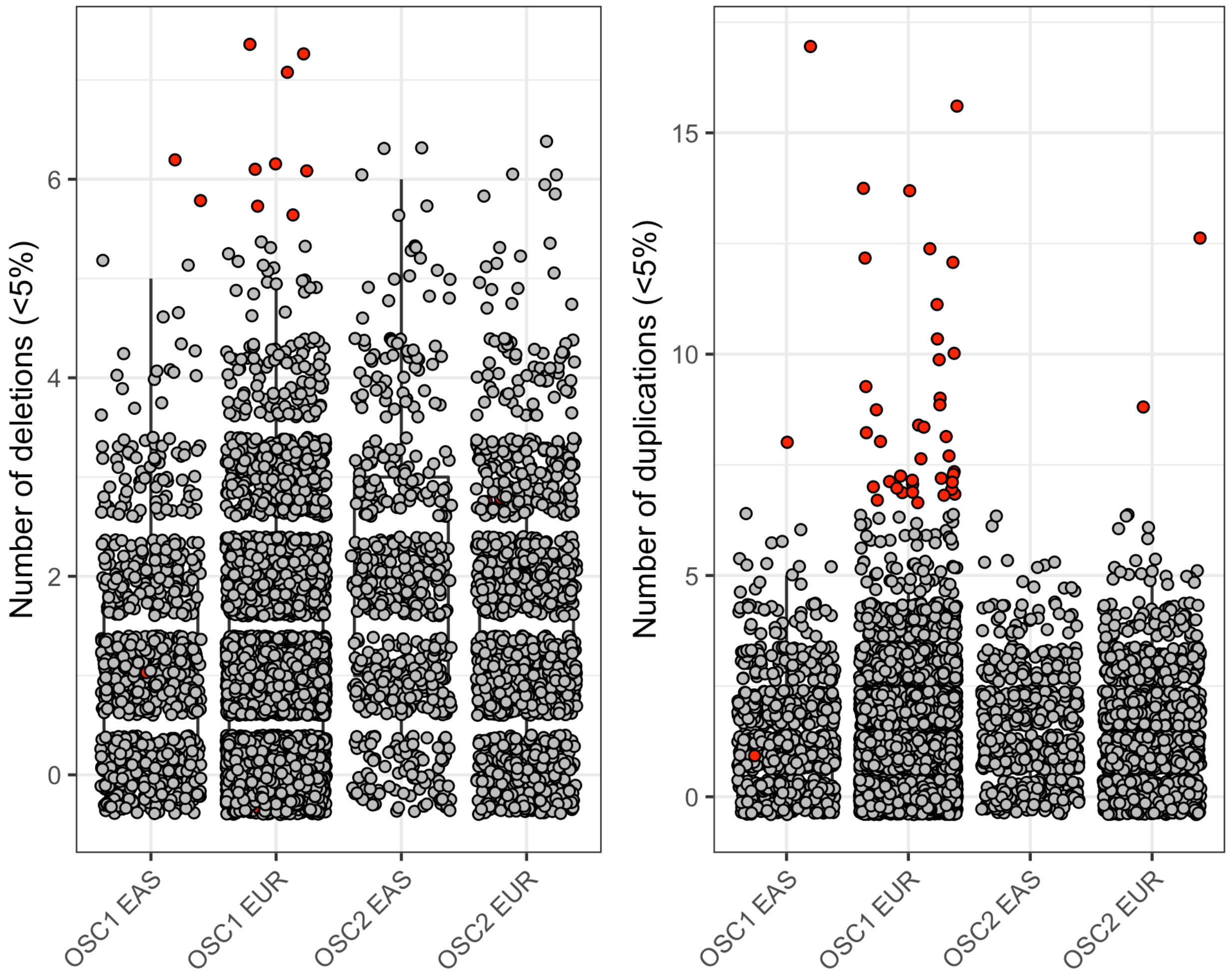


**Fig. S1:** The distribution of the number of CNVs (<5%) per individual stratified by study phases and population groups. Red circles indicate samples with excessive deletions or duplications.

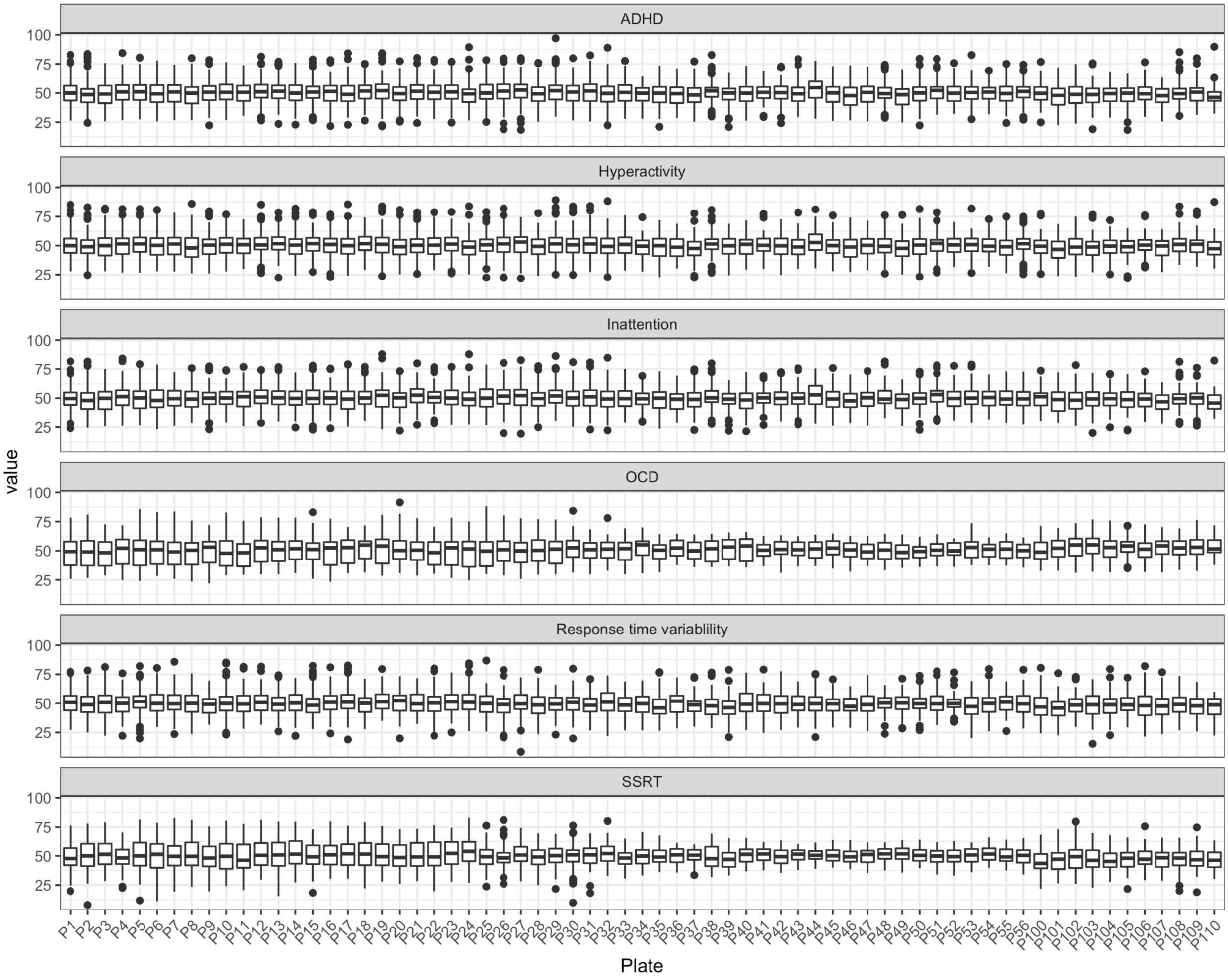


**Fig. S2**: The distribution of T-scores stratified by genotyping plates. The six panels are for six different trait T-scores. Boxplots show median, Q1 and Q3. Their arms indicate 1.5IQR boundary and dots are the samples with T-scores beyond the 1.5IQR boundary.

**w**

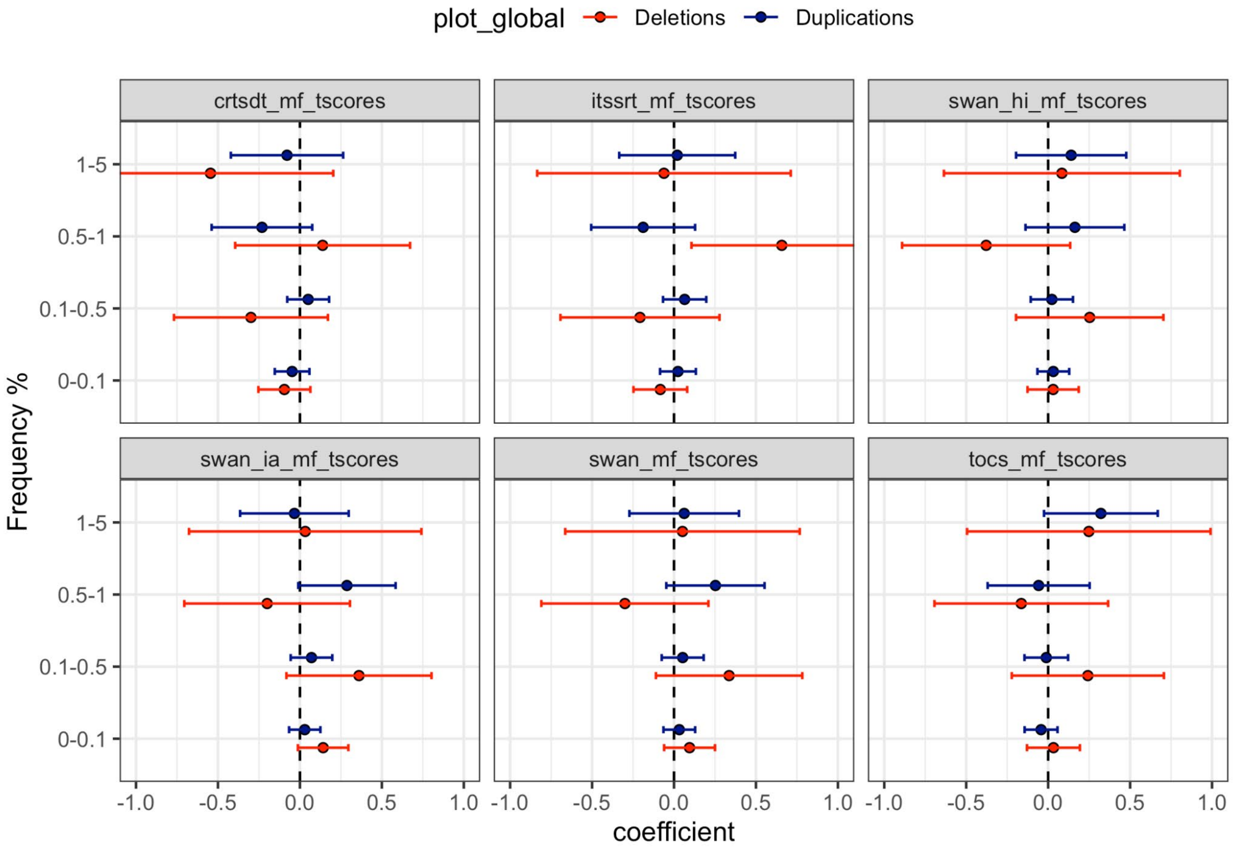


**Fig. S3**: Global burden analysis of CNVs within different frequency bins. The middle points show the beta coefficient and the error bars show the 95% confidence intervals of the coefficient from linear regression model regressing the number of genes impacted by CNVs per individual on the T-score. Different colors denote deletions (red) and duplications (blue).

**Supplementary Tables:**

**Supplementary Table S1:** A selection of publications on clinically relevant CNVs/ associations in mental health illnesses [see online Supplementary Table S1.xlsx]

**Supplementary Table S2:** Participant information, phenotypes distribution, CNV calls, rare loss-of-function, and rare predicted damaging missense variants from the Spit for Science cohort used in the current study [see online Supplementary Table S2.xlsx]

**Supplementary Table S3:** Clinically relevant variants in Spit for Science cohort. [see online Supplementary Table_S3.xlsx].

**Supplementary Table S4:** Clinically relevant variants in ALSPAC cohort. [see online Supplementary Table_S4.xlsx].

**Supplementary Table S5:** Burden findings and loci association results in Spit for Science cohort. [see online Supplementary Table_S5.xlsx].

**Supplementary Table S6:** Comparison among various general population CNV studies [see online Supplementary Table_S6.xlsx].

**Acknowledgments:**

We are extremely grateful to all the families who took part in this study, the midwives for their help in recruiting them, and the whole ALSPAC team, which includes interviewers, computer and laboratory technicians, clerical workers, research scientists, volunteers, managers, receptionists and nurses. The UK Medical Research Council and Wellcome (Grant ref: 217065/Z/19/Z) and the University of Bristol provide core support for ALSPAC. This publication is the work of the authors and Marc Woodbury-Smith will serve as guarantors for the contents of this paper.

Funding support for the Study of Addiction: Genetics and Environment (SAGE) was provided through the NIH Genes, Environment and Health Initiative [GEI] (U01 HG004422). SAGE is one of the genome-wide association studies funded as part of the Gene Environment Association Studies (GENEVA) under GEI. Assistance with phenotype harmonization and genotype cleaning, as well as with general study coordination, was provided by the GENEVA Coordinating Center (U01 HG004446). Assistance with data cleaning was provided by the National Center for Biotechnology Information. Support for collection of datasets and samples was provided by the Collaborative Study on the Genetics of Alcoholism (COGA; U10 AA008401), the Collaborative Genetic Study of Nicotine Dependence (COGEND; P01 CA089392), and the Family Study of Cocaine Dependence (FSCD; R01 DA013423). Funding support for genotyping, which was performed at the Johns Hopkins University Center for Inherited Disease Research, was provided by the NIH GEI (U01HG004438), the National Institute on Alcohol Abuse and Alcoholism, the National Institute on Drug Abuse, and the NIH contract "High throughput genotyping for studying the genetic contributions to human disease" (HHSN268200782096C). The datasets used for the analyses described in this manuscript were obtained from dbGaP at http://www.ncbi.nlm.nih.gov/projects/gap/cgi-bin/study.cgi?study_id=phs000092.v1.p1 through dbGaP accession number phs000092.v1.p1.

The authors acknowledge the contribution of data from Genetic Architecture of Smoking and Smoking Cessation accessed through dbGaP. Funding support for genotyping, which was performed at the Center for Inherited Disease Research (CIDR), was provided by 1 X01 HG005274-01. CIDR is fully funded through a federal contract from the National Institutes of Health to The Johns Hopkins University, contract number HHSN268200782096C. Assistance with genotype cleaning, as well as with general study coordination, was provided by the Gene Environment Association Studies (GENEVA) Coordinating Center (U01 HG004446). Funding support for collection of datasets and samples was provided by the Collaborative Genetic Study of Nicotine Dependence (COGEND; P01 CA089392) and the University of Wisconsin Transdisciplinary Tobacco Use Research Center (P50 DA019706, P50 CA084724). The datasets used for the analyses described in this manuscript were obtained from dbGaP at https://www.ncbi.nlm.nih.gov/projects/gap/cgi-bin/study.cgi?study_id=phs000404.v1.p1 through dbGaP accession number phs000404.v1.p1.

The dataset(s) used for the analyses described in this manuscript were obtained from the NEI Refractive Error Collaboration (NEIREC). Funding support for NEIREC was provided by the National Eye Institute. We would like to thank NEIREC participants and the NEIREC Research Group for their valuable contribution to this research. The datasets used for the analyses described in this manuscript were obtained from dbGaP at https://www.ncbi.nlm.nih.gov/projects/gap/cgi-bin/study.cgi?study_id=phs000303.v1.p1 through dbGaP accession number phs000303.v1.p1.

Funding support for the "CIDR Visceral Adiposity Study" was provided through the Division of Aging Biology and the Division of Geriatrics and Clinical Gerontology, NIA. The CIDR Visceral Adiposity Study includes a genome-wide association study funded as part of the Division of Aging Biology and the Division of Geriatrics and Clinical Gerontology, NIA. Assistance with phenotype harmonization and genotype cleaning, as well as with general study coordination, was provided by Heath ABC Study Investigators. The datasets used for the analyses described in this manuscript were obtained from dbGaP at https://www.ncbi.nlm.nih.gov/projects/gap/cgi-bin/study.cgi?study_id=phs000169.v1.p1 through dbGaP accession number phs000169.v1.p1.
